# Cohort profile: COVID-19 in a cohort of pregnant women and their descendants, the MOACC-19 study

**DOI:** 10.1101/2020.08.20.20178657

**Authors:** Javier Llorca, Carolina Lechosa-Muñiz, Pilar Gortázar, María Fernández-Ortiz, Yolanda Jubete, María J. Cabero, the MOACC-19 group, Jéssica Alonso-Molero, Bárbara Arozamena, Laura Conde-Gil, Elsa Cornejo del Río, Rocío Cuesta-González, Trinidad Dierssen-Sotos, Pelayo Frank de Zulueta, Inés Gómez-Acebo, Coral Llano-Ruiz, Lorena Lasarte-Oria, Sonia López-Gómez, Sonia Mateo-Sota, Victoria Orallo, Rosa Pardo, Daniel Pérez González, María Sáez de Adana Herrero

**Author notes:** Correspondence: Javier Llorca, Facultad de Medicina, Universidad de Cantabria, Avda. Herrera Oria s/n, 39011 Santander, Spain.

## Abstract

**Purpose:** The Mother and Child Covid-19 study is a cohort recruiting pregnant women and their children in Cantabria, North of Spain, during COVID-19 pandemic in order to ascertain Consequences of SARS-CoV-2 infection on pregnant women and their descendants. This article reports the cohort profile and preliminary results as recruitment is still open.

**Participants:** Three sub-cohorts can be identified at recruitment. Sub-cohort 1 includes women giving birth between 23^rd^ March and 25^th^ May 2020; they have been retrospectively recruited and could have been exposed to COVID-19 only in their third trimester of pregnancy. Sub-cohort 2 includes women giving birth from 26^th^ May 2020 on; they are being prospectively recruited and could have been exposed to COVID-19 in both their second and third trimesters of pregnancy. Sub-cohort 3 includes women in their 12^th^ week of pregnancy prospectively recruited from 26^th^ May 2020 on; they could have been exposed to COVID-19 anytime in their pregnancy. All women are being tested for SARS-CoV-2 infection using both RT-PCR for RNA detection and ELISA for anti-SARS-CoV-2 antibodies. All neonates are being tested for antibodies using immunochemoluminiscency tests; if the mother is tested positive for SARS-CoV-2 RNA, a naso-pharyngeal swab is also obtained from the child for RT-PCR analysis.

**Findings to date:** As of 22^nd^ October, 1167 women have been recruited (266, 354 and 547 for sub-cohorts 1, 2 and 3, respectively). Fourteen women tested positive to SARS-CoV-2 RNA by the day of delivery. All fourteen children born from these women tested negative for SARS-CoV-2 RNA.

**Future plans:** Children from women included in sub-cohort 3 are expected to be recruited by the end of 2020. Children will be followed-up for one year in order to ascertain the effect that COVID-19 on their development.

**ARTICLE SUMMARY:** Strengths and limitations

**Strengths:**

- This cohort would ascertain the effect of COVID-19 in both mother and children whatever the trimester of the infection.
- It would also compare health care provided to pregnant women during the COVID-19 pandemic with that provided in the same hospital before the emergence of COVID-19.
- The cohort is recruited in Spain, one of the developed countries earlier and more affected by COVID-19.

**Limitations:**

- The study could be underpowered according to the prevalence reported in a Spanish national study.
- Information regarding exposure to people infected by SARS-CoV-2 or risk activities is self-reported.

## INTRODUCTION

The emergence of the new coronavirus SARS-CoV-2 in China at the end of 2019 produced a pandemic of COVID-19 characterized by fever, cough, pneumonia and other respiratory symptoms, with many patients also developing a systemic inflammatory crisis, sometimes considered a cytokine storm (1,2). Both bilateral pneumonia and cytokine storm could eventually lead to severe disease, especially in vulnerable groups, reaching a case-fatality rate about 3% (2).

As the number of COVID-19 cases increases, concern arises on the role played by pregnant women, whether as vulnerable group or as putative transmitters to their descendant (3). Two other coronaviruses had produced epidemics of international interest in the 21^st^ century. During the Severe Acute Respiratory Syndrome (SARS) epidemic in 2002-2003, infection in pregnancy was associated with severe maternal disease, maternal mortality and spontaneous miscarriage (4). The Middle-East Respiratory Syndrome (MERS) appeared in 2012 and it is still ongoing. Only 11 cases of MERS in pregnancy have been reported, ten of them having adverse clinical outcome (3). Vertical transmission has not been documented in either SARS or MERS (3).Initial reports indicated that most infected women have mild presentation (5-7) and maternal mortality in COVID-19 pregnant women is scarce as compared with both SARS and MERS (8). Nevertheless, pregnant women are more likely to be admitted to ICU (9-11) and suffer postpartum complications (12) than non-pregnant women of similar age. Possible mechanisms for bringing about worse health outcome in pregnant women could encompass changes in lung volume, increased secretions in the upper respiratory tract, increasing susceptibility due to changes in cell-mediated immunity (8,13).

While two articles have reported remarkable decreases in newborns with gestational age lower than 28 weeks to non-SARS-CoV-2 infected women during the pandemic (14,15), high rates of preterm delivery by caesarean rate have been reported in COVID-19 infected women (7,10,12). No differences in caesarean rate delivery were found, however, when comparing symptomatic vs. asymptomatic infected women (12), which suggests that the medical ground for COVID-19 associated caesarean rates was the infection itself rather than the clinical situation.

Putative ways of mother to child transmission of COVID-19 to be considered include placental, intravaginal or breastfeeding transmissions. Reports on neonate outcome from women infected by SARS-CoV-2 are still scarce. Although most articles did not find evidence of vertical transmission (16-19), some cases of infected newborns have been documented in spite of delivering by caesarean rate, avoiding breastfeeding and careful mother - child isolation (10,20–23). The case for transplacental transmission, however, is subject to stringent requisites, including the detection of SARS-CoV-2 by PCR in umbilical cord blood, neonatal blood collected within the first 12 hours of life or amniotic fluid collected prior to rupture of membrane (24). In this regard, Vivanti et al (25) have convincingly reported a well-documented case of transplacental transmission. Placental pathology such us thrombi in foetal vessels has been found to be frequent in infected pregnant women (12) and could be a way of damaging neonates even in absence of SARS-CoV-2 mother-to-child transmission.

Besides the importance that COVID-19 disease could directly have on pregnant women and pregnancy result, there is also an indirect way that has not been studied in deep so far. Many countries have fought against COVID-19 by locking down most economic and social activity (26) and deeply changing the way hospitals were working (e.g., many consultations were carried out via phone or other non-face-to-face technologies; surgical procedures were delayed) (27), leading to noticeable changes in emergency room motives of consultation, even with strong decreases in consultations by usually urgent and life-threatening diseases (28,29) so that the whole health system in many countries has been deflected with unpredictable consequences. There is still no data on whether this switch could have affected the way that pregnant women with or without COVID-19 have been attended during pregnancy and delivery. For instance, we may wonder if caesarean rates could have rose or if the number of consultations during pregnancy could have decreased.

Finally, some studies have reported higher risk of COVID-19 in the most deprived (30) or with lower educational attainment (31). In pregnant women, however, the impact of socio-economic status on SARS-CoV-2 infection is still little known and only ecological studies with small sample size have been published (32).

In this article, we are reporting the inception of a cohort of pregnant women and their neonates during the COVID-19 pandemic in Spain, one of the European countries most relentlessly stroke by COVID-19. Our main goals are to ascertain differences in outcomes between pregnant women with and without COVID-19, as well as among their children, and to compare pregnancy outcomes during COVID-19 pandemic with those occurred in a pre-COVID-19 cohort in the same hospital (33, 34).

## METHODS

### Specific and broader aims and rationale of study design

This article aims to report the design, implementation and early results of the MOther And Child Covid-19 cohort (MOACC-19) incepted at the University Hospital Marqués de Valdecilla (HUMV), Santander, Spain.

The broader aim of MOACC-19 is to better understand the effect that COVID-19 pandemic has on both mother and child health. The specific objectives are: (1) To estimate the prevalence of anti-SARS-CoV-2 antibodies in pregnant women; (2) to ascertain the risk of vertical transmission; (3) to find out the impact of both symptomatic and asymptomatic infection of mother on child health at delivery and after 6 and 12 months of follow-up; (4) to evaluate the modifications in medical practice in pregnancy, delivery and neonatal care during COVID-19 pandemic, as well as the changes they could have on neonate health, and (5) to evaluate the relationship between socio-economic status and risk of infection by SARS-CoV-2 in pregnant women.

### Context: Covid-19 pandemic in Spain

As of 13^th^ December, COVID-19 has produced 1.73 million cases in Spain (cumulative incidence: 3682 per 100000 people) and has claimed for 47624 deaths (mortality rate: 101.3 per 100000 people) according to official data reported to ECDC (35). Daily number of cases are displayed in Figure 1. The first case was reported by 2nd February and the first death by 5th March. The daily number of cases in the first wave peaked by 27th March (9181 cases) and that of deaths by 3rd April (950 deaths). Figure 1 also presents the main legal restrictions ordered by the Spanish Government, including severe confinement from 27th March to 21st June and complete lockdown from 29th of March to 12th of April.

**Figure 1.**
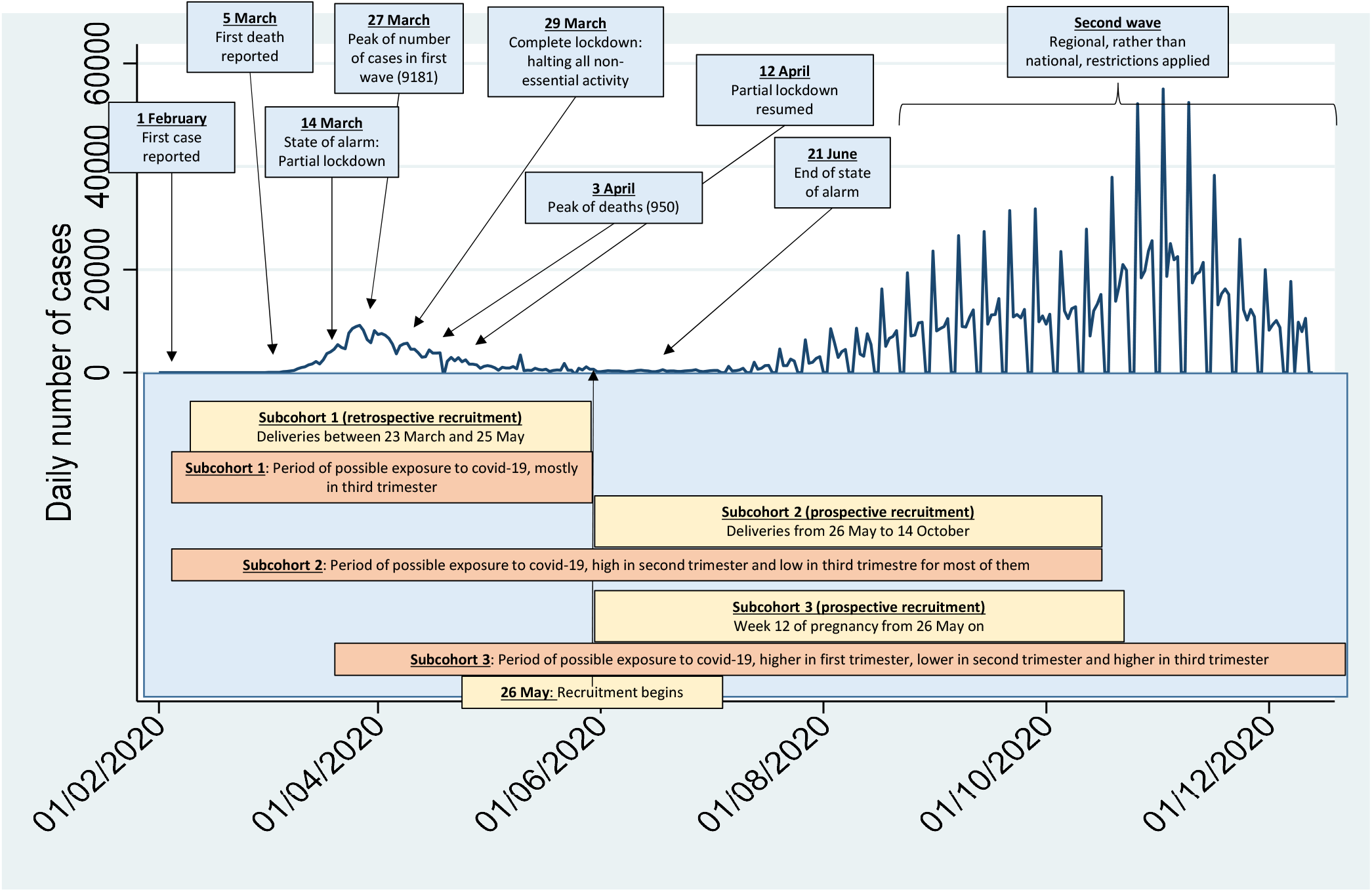
Pregnant women recruitment for this study in the context of the COVID-19 pandemic evolution in Spain. The blue line represents the daily number of cases as reported to the ECDC up to 13^th^ December, 2020 (reference # 35). The blue rectangles indicate the main legal restrictions. The yellow rectangles mark the recruitment period for each sub-cohort. The orange rectangles indicate the period of exposure to COVID-19 for each sub-cohort. Note on reported cases: Peaks by 10^th^, 22^nd^ and 28^th^ May, and troughs by 19^th^ April and 5^th^ May are anomalies in the number of reported cases due to corrections in the series. From 1^st^ July on, cases are not reported on Saturdays and Sundays, so number of cases on Mondays represent three-day cumulative figures.

### Setting

The HUMV is a third level hospital with 900 beds located in the region of Cantabria, North of Spain. It usually attends about 2500 deliveries in a normal year (about 90% deliveries in the region), but due to the COVID-19 crisis all deliveries occurred in the Cantabria region from March to June 2020 have been gathered in the HUMV. From March to June 2020, it also concentrated all COVID-19 admissions in Cantabria; in order to do it, the hospital was divided in two separated parts, one for COVID-19 patients and the other for non-COVID-19 patients. From 23^th^ March 2020 on, all pregnant women admitted for delivery were tested to SARS-CoV-2 active infection using RT-PCR.

### Recruitment

MOACC-19 is intended to recruit at least 1000 pregnant women and their neonates. Recruitment timing is displayed in Figure 1 in the context of the pandemic evolution in Spain. Recruitment begun on 26^th^ May 2020. It was organized with three sub-cohorts in mind:

– Sub-cohort 1: Women who had delivered from 23^th^ March to 25^th^ May 2020. They had already been tested with RT-PCR by the day of delivery. They are being retrospectively contacted by phone and invited to participate in MOACC-19. These women had been exposed to SARS-CoV-2 in the third trimester of their pregnancy.
– Sub-cohort 2: Women admitted to delivery from 26^th^ May to 14^th^ October 2020 were invited to participate in the study at admission. They could have been exposed to SARS-CoV-2 in the second –where the pandemic in Spain was higher- or third trimester of their pregnancy. As most of them delivered a baby before the irruption of the second wave, the later their date of delivery, the lower their exposure to COVID-19 in the third trimester.
– Sub-cohort 3: Women consulting for their 12^th^ week of pregnancy from 26^th^ May on are being invited to participate. If they agreed, they are immediately tested with RT-PCR. They have been exposed to the worst period of the pandemic in their first trimester; their exposure in the second trimester was lower as the pandemic first wave went down, but their exposure in the third trimester is increasing as the second wave develops.

Women in sub-cohorts 1 and 2 are also invited to include their neonates in the study. Women in sub-cohort 3 will be invited to do so by the time of delivery.

Figure 2 displays the flow diagrams of tasks carried out for each sub-cohort, including data collection, biological samples, biological determinations and follow up.

**Figure 2.**
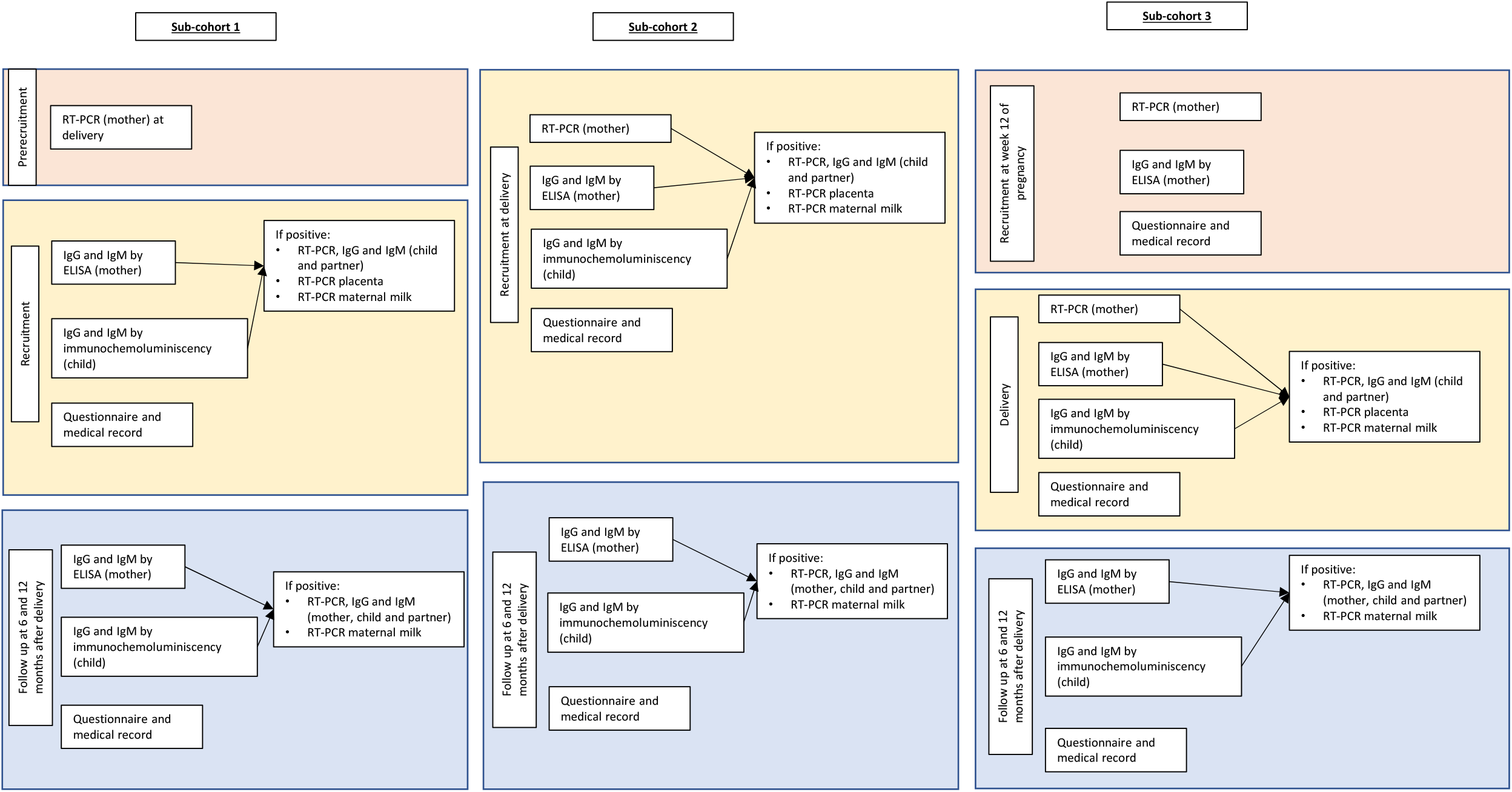
Flow diagram of tasks performed in each sub-cohort

### Data collection

At recruitment, women are being asked to answer a face-to-face questionnaire. It included socio-demographic data, obstetrics history, medical history, exposure to COVID-19 and symptoms compatible with COVID-19. Data on both obstetrics and medical history are to be completed by reviewing medical records. Regarding the neonate, we will review medical records in order to obtain information on characteristics at birth, perinatal pathology, admission to neonatal ICU and type of feeding at hospital discharge (Table 1).

**Table 1.**
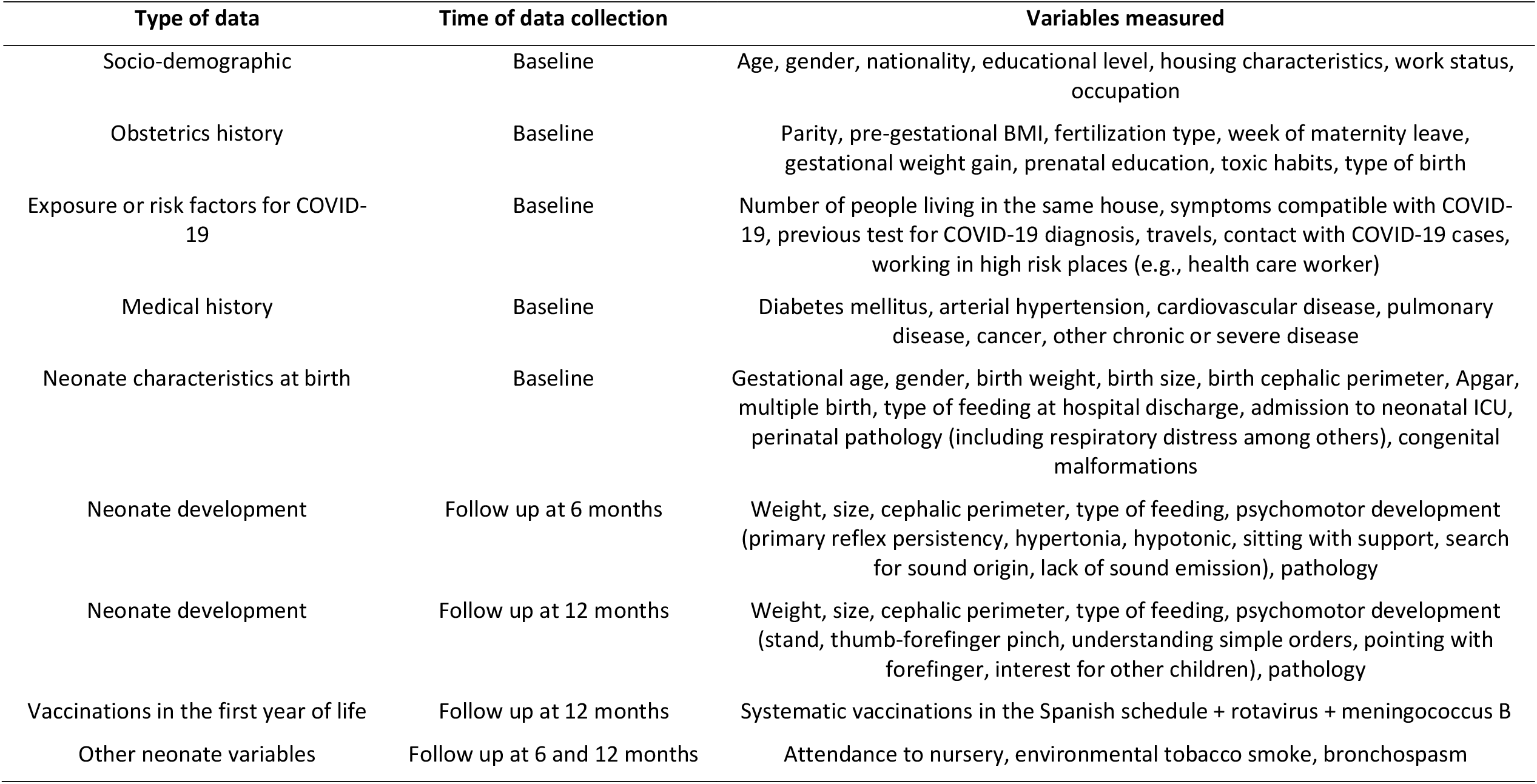
Summary of data collected in the MOACC-19 study

### Follow up

Neonates will be followed up at 6 and 12 months of life. They will be explored by a paediatrician in order to ascertain their general development -via general exploration- and their psychomotor development -via Denver Developmental Screening Test (DDST-II)- (Table 1). Type of feeding, vaccinations, exposure to environmental tobacco smoke, respiratory diseases and other diseases in the first year of life will be asked for (Table 1).

### Biological determinations

A naso-pharyngeal sample is being taken with a swab from all women at delivery and from women in sub-cohort 3 at 12th week of pregnancy. These samples are being tested for SARS-CoV-2 infection via RT-PCR. A blood sample by venopuncture is being obtained from each woman at recruitment and tested for anti-SARS-CoV-2 spike protein IgG and IgM using ELISA. Had any of these determinations in the mother been positive, we would carry out both RT-PCR and antibody determinations via ELISA for the neonate and the woman’s partner. Moreover, we would search for viral RNA in placenta and mother’s milk via RT-PCR.

Each neonate in Spain is screened for congenital metabolopathies by obtaining a blood sample from the heel in the first few days of life. In order to avoid unnecessary pricks to a neonate, in the same procedure we are obtaining a blood gout for studying IgG and IgM via immunochemoluminiscency.

In the follow up, new blood samples by venopuncture (mother) or finger prick (child) will be obtained at 6 and 12 months in order to study anti-SARS-CoV-2 antibodies.

### Comparison group

Health care during pregnancy and at delivery, newborn characteristics, development and non-COVID-19 pathology will be compared with a cohort of 969 neonates recruited in 2018 in the same hospital. This cohort has been described elsewhere (34,35). In brief, the cohort was recruited from January to August 2018 in the HUMV; data on maternal age, parity, educational level, pregnancy duration, type of delivery and toxic habits in pregnancy were obtained from maternal medical records. Data on neonate gender, weight and other characteristics at birth, attendance to nursery and type of feeding were obtained by interviewing the mother in each check-up every other month until the 12th month of life. Data on newborn health evolution, including vaccinations, infectious diseases, bronchospasms, emergency room consultations and hospital admissions were obtained from medical records.

### Statistical analysis conducted to date

Descriptive statistics are displayed as frequency and percentage for categorical variables and mean and standard deviation for continuous variables. Statistical comparisons are performed via chi-squared test or independent samples Student-t test.

### Ongoing statistical analysis

In this section we outline the ongoing statistical analysis for each specific objective.

1. Prevalence and its 95% CI of anti-SARS-CoV-2 antibodies in pregnant women will be estimated assuming a binomial distribution (or a Poisson distribution if the number of positives is too small).
2. Risk of vertical transmission would be described. We do not expect that the number of mother and children positive to SARS-CoV-2 either antibodies or RNA would be enough to perform a formal statistical analysis.
3. The impact of maternal COVID-19 on child health will be evaluated using ANCOVA for continuous effect variables (e.g., weight and cephalic perimeter at birth) or logistic regression for dichotomic effect variables (e.g., premature birth, respiratory distress). Both ANCOVA and logistic regression models will be adjusted for the identifiable confounders.
4. In order to evaluate the modifications in medical practice in pregnancy we will compare the MOACC-19 cohort with that recruited in 2018 in the same hospital. Continuous variables will be studied using Student t test for independent samples and ANCOVA in order to adjust for confounding variables. Categorical variables will be studied using chi-squared test. The effect those modifications in medical practice could have on child health will be studied with the same statistical techniques indicated in specific objective 3. This comparison will be made once all children have been recruited.
5. Socio-economic status will be measured using HOUSES, a score developed by Jung et al. It includes four data from housing: surface, cost, number of restrooms and number of bedrooms. Its relationship with risk of infection by SARS-CoV-2 in pregnant women will be analysed via logistic regression.

### Patient and public involvement

Patients were not involved in the study.

### Ethical considerations

The study was approved by the Clinical Research Ethics Committee of Cantabria (reference: 2020.174). Two different written informed consents -one for the mother and one for the child-have to be signed by the mother before being admitted in the study. The study is conducted according to the Declaration of Helsinki (last update of Fortaleza) and the European Union regulation 2016/679 on the protection of natural persons with regard to the processing of personal data.

## RESULTS AND DISCUSSION

### Findings to date

As of 22^nd^ October, where women recruitment was closed, 1167 women have been recruited; 266 delivered before 26^th^ May and were recruited retrospectively; 354 delivered after 26^th^ May and were recruited prospectively and 547 reached their 12^th^ week of pregnancy after 26^th^ May. Their characteristics appear in Table 2. About 50% were 35 year or older, 88% were European and 10% were born in Latino-America. 47% reported university studies, 75% were actively working. 9% pregnancies were produced by in vitro fertilization or artificial insemination. 7.3% women reported to have smoked in pregnancy, but none among those recruited while still pregnant (p<0.001); 2.4% women reported to have drunk alcohol in pregnancy, but none among those still pregnant (p < 0.001). For 54% women, this was their first child. Caesarean rate was carried out in 18% deliveries and other instrumental procedures in 7%. The three sub-cohorts recruited differed in educational level (women in sub-cohort 1 reached less likely university studies) and smoking in pregnancy (0% reported by women in sub-cohort 3, p<0.001).

**Table 2.** Main characteristics of women included in MOACC-19 study

| Variable | Whole cohort<br>(n = 1167) | Sub-cohort 1<br>(women delivering before<br>26/May/2020)<br>(n = 266) | Sub-cohort 2<br>(women delivering from<br>26/May/2020 on)<br>(n = 354) | Sub-cohort 3<br>(women in their 12th week of pregnancy from<br>26/May/2020)<br>(n = 547) | P |
| --- | --- | --- | --- | --- | --- |
| Age, mean±sd | 34.0±5.0 | 34.4±4.8 | 33.8±5.2 | 34.0±5.0 | 0.38 |
| Age |  |  |  |  |  |
| <25 | 45 (3.9) | 8 (3.0) | 18 (5.1) | 19 (3.5) | 0.71 |
| 25-29 | 161 (13.8) | 33 (12.4) | 53 (15.1) | 75 (13.7) |  |
| 30-34 | 377 (32.4) | 85 (32.0) | 114 (32.4) | 178 (32.5) |  |
| 35-39 | 436 (37.4) | 106 (39.9) | 119 (33.8) | 211 (38.6) |  |
| >40 | 146 (12.5) | 34 (12.8) | 48 (13.6) | 64 (11.7) |  |
| Nationality |  |  |  |  |  |
| European | 1014 (87.7) | 241 (90.9) | 303 (87.3) | 470 (86.4) | 0.43 |
| African | 18 (1.6) | 2 (0.8) | 8 (2.3) | 8 (1.5) |  |
| Asiatic | 7 (0.6) | 1 (0.4) | 3 (0.9) | 3 (0.6) |  |
| Latino-America | 117 (10.1) | 21 (7.9) | 33 (9.5) | 63 (11.6) |  |
| Education level |  |  |  |  |  |
| Primary | 175 (15.1) | 33 (12.4) | 50 (14.4) | 92 (16.9) | 0.007 |
| Secondary | 73 (6.3) | 18 (6.8) | 34 (9.8) | 21 (3.9) |  |
| Vocational training | 369 (31.8) | 99 (37.2) | 98 (28.2) | 172 (31.5) |  |
| University | 543 (46.8) | 116 (43.6) | 166 (47.7) | 261 (47.8) |  |
| Working status |  |  |  |  |  |
| Unemployed / no active worker | 277 (23.9) | 56 (21.2) | 89 (25.5) | 132 (24.2) | 0.55 |
| Employed | 864 (74.6) | 205 (77.7) | 256 (73.4) | 403 (73.8) |  |
| Student | 18 (1.6) | 3 (1.1) | 4 (1.2) | 11 (2.0) |  |
| Gestational age leave work, mean±sd | 24.3±9.8 | 25.3±9.3 | 23.5±10.1 | - | 0.06 |
| Fertilization type |  |  |  |  |  |
| Natural | 557 (91.5) | 245 (92.8) | 312 (90.4) | - | 0.42 |
| Artificial insemination | 8 (1.3) | 2 (0.8) | 6 (1.7) | - |  |
| In vitro fertilization (own ovules) | 32 (5.3) | 14 (5.3) | 18 (5.2) | - |  |
| In vitro fertilization (donated ovules) | 12 (2.0) | 3 (1.1) | 9 (2.6) | - |  |
| Pregestational BMI, mean±sd | 24.4±4.8 | 24.3±4.8 | 24.3±5.1 | 24.5±4.6 | 0.74 |
| Gestational weight gain, mean±sd | 12.1±5.1 | 11.9±5.4 | 12.3±4.9 | - | 0.29 |
| Smoker in pregnancy | 85 (7.3) | 31 (11.7) | 54 (15.4) | 0 (0.0) | <0.001 |
| Alcohol consumption in pregnancy | 28 (2.4) | 13 (4.9) | 15 (4.3) | 0 (0.0) | <0.001 |
| Parity (incl. current delivery) |  |  |  |  |  |
| 1 | 335 (54.4) | 132 (49.6) | 203 (58.0) | - | 0.23 |
| 2 | 235 (38.2) | 114 (42.9) | 121 (34.6) | - |  |
| ≥3 | 46 (7.5) | 20 (7.5) | 26 (7.4) | - |  |
| Type of delivery |  |  |  |  |  |
| Eutocic | 456 (75.0) | 195 (73.9) | 261 (75.9) | - | 0.81 |
| Instrumental | 40 (6.6) | 19 (7.2) | 21 (6.1) | - |  |
| Caesarean rate | 112 (18.4) | 50 (18.9) | 62 (18.0) | - |  |
| COVID-19 RT-PCR<br>(women) |  |  |  |  |  |
| Positive | 14 (2.3) | 5 (1.9) | 9 (2.6) | - | 0.56 |
| Negative | 600 (97.7) | 261 (98.1) | 339 (97.4) | - |  |
| COVID-19 RT-PCR<br>(partner) |  |  |  |  |  |
| Positive | 12 (2.0) | 1 (0.4) | 11 (3.2) | - | 0.02 |
| Negative | 575 (98.0) | 247 (99.6) | 328 (96.8) | - |  |
| Pathology in pregnancy | 409 (65.3) | 178 (66.9) | 231 (65.3) | - | 0.67 |
| Gestational diabetes | 46 (7.4) | 18 (6.8) | 28 (7.9) | - | 0.59 |
| Gestational diabetes with insulin | 26 (4.2) | 13 (4.9) | 13 (3.7) | - | 0.46 |
| Gestational hypertension | 22 (3.6) | 9 (3.4) | 13 (3.7) | - | 0.85 |
| Chronic hypertension | 5 (0.8) | 2 (0.8) | 3 (0.9) | - | 0.90 |
| Preeclampsia | 24 (3.9) | 9 (3.4) | 15 (4.2) | - | 0.59 |
| Placenta previa | 0 | 0 | 0 | - | - |
| Placental abruption | 6 (1.0) | 3 (1.1) | 3 (0.9) | - | 0.72 |
| Threatened miscarriage | 26 (4.2) | 14 (5.3) | 12 (3.4) | - | 0.25 |
| Metrorrhagia (2<sup>nd</sup> half pregnancy) | 5 (0.8) | 3 (1.1) | 2 (0.6) | - | 0.44 |
| Prelabour rupture of membranes | 3 (0.5) | 1 (0.4) | 2 (0.6) | - | 0.74 |
| Intrahepatic cholestasis | 4 (0.7) | 3 (1.1) | 1 (0.3) | - | 0.19 |
| Oligoamnios | 3 (0.5) | 3 (1.1) | 0 | - | 0.05 |
| Polyamnios | 0 | 0 | 0 | - | - |
| Surgery | 10 (1.6) | 6 (2.3) | 4 (1.1) | - | 0.27 |
| Threatened premature delivery | 12 (1.9) | 5 (1.9) | 7 (2.0) | - | 0.93 |
| Chorioamnionitis | 1 (0.2) | 0 | 1 (0.3) | - | 0.39 |
| Chromosomopathies | 3 (0.5) | 3 (1.1) | 0 | - | 0.05 |

Fourteen women in sub-cohorts 1 and 2 tested positive to SARS-CoV-2 RNA via RT-PCR; five of them were from the retrospective sample, and 9 from the prospective sample (Supplementary Table 1). Seven of their partners were also positive to RT-PCR and 7 were negative. Seven women reported university studies; nine were employed (4 as health care workers and 2 worked in restaurants or commerce). Caesarean rate was performed in 4 women and other instrumental delivery in other 2. They did not suffer any gestational pathology other than COVID-19. Six were asymptomatic. A woman developed shortness of breathing and suffered syncope by week 32^nd^ of pregnancy; by week 39^th^ she delivered a healthy child. Seven women reported to have had contact at home with someone diagnosed of COVID-19 and another with infected relative or friends; the remaining women reported neither a known contact responsible for them to get infected nor an international travel in the previous 2 weeks.

Six hundred and thirteen children had been born from sub-cohorts 1 and 2. Their main characteristics appear in Table 3. Twenty-seven of them (4.5%) were premature; 35 children (5.7%) had low weight at birth and 26 (4.2%) weighted more than 4000 g. Eight children were twins (1.3%). Fourty children (6.5%) required admission to ICU; 10 because of jaundice, 9 due to respiratory distress and 5 due to low weight at birth or prematurity. About 60% children were exclusively breastfeeding at hospital discharge, 22% received mixed feeding and 18% were fed with infant formula. Child characteristics were similar in children retrospectively or prospectively recruited.

**Table 3.** Main characteristics of children included in the study

| Variable | Total<br>(n = 613) | Sub-cohort 1<br>(children born<br>before<br>26/May/2020)<br>(n = 266) | Sub-cohort 2<br>(children born<br>from<br>26/May/2020 on)<br>(n = 347) | P value |
| --- | --- | --- | --- | --- |
| **Gestational age at delivery** |  |  |  |  |
| <34 weeks | 6 (1.0) | 4 (1.5) | 2 (0.6) | 0.50 |
| 34-36 weeks | 21 (3.5) | 9 (3.5) | 12 (3.5) |  |
| ≥37 weeks | 576 (95.5) | 247 (95.0) | 329 (95.9) |  |
| **Birth weight** |  |  |  |  |
| <2500 g | 35 (5.7) | 17 (6.4) | 18 (5.2) | 0.63 |
| 2500 – 4000 g | 552 (90.1) | 236 (88.7) | 316 (91.1) |  |
| >4000 g | 26 (4.2) | 13 (4.9) | 13 (3.8) |  |
| **Birth size, mean±sd** | 49.7±2.6 | 49.4±2.9 | 50.0±2.3 | 0.01 |
| **Birth cephalic perimeter, mean±sd** | 34.4±1.4 | 34.3±1.6 | 34.5±1.3 | 0.09 |
| **Apgar 1', mean±sd** | 8.7±0.8 | 8.7±0.8 | 8.7±0.8 | 0.55 |
| **Apgar 5', mean±sd** | 9.7±0.6 | 9.7±0.6 | 9.6±0.6 | 0.29 |
| **pH, mean±sd** | 7.24±0.31 | 7.23±0.47 | 7.26±0.08 | 0.31 |
| **Gender** |  |  |  |  |
| Woman | 295 (48.4) | 119 (45.4) | 176 (50.7) | 0.20 |
| Man | 314 (51.6) | 143 (54.6) | 171 (49.3) |  |
| **Twin** | 8 (1.3) | 2 (0.8) | 6 (1.7) | 0.30 |
| **Type of feeding at hospital discharge** |  |  |  |  |
| Exclusive breastfeeding | 360 (60.2) | 156 (59.8) | 204 (60.5) | 0.67 |
| Mixed | 130 (21.7) | 54 (20.7) | 76 (22.6) |  |
| Infant formula | 108 (18.1) | 51 (19.5) | 57 (16.9) |  |
| **Neonatology admission** | 40 (6.5) | 23 (8.7) | 17 (4.9) | 0.06 |
| Jaundice | 10 (1.6) | 7 (2.6) | 3 (0.9) | 0.08 |
| Respiratory distress | 9 (1.5) | 5 (1.9) | 4 (1.1) | 0.44 |
| Low weight | 1 (0.2) | 1 (0.4) | 0 (0.0) | 0.25 |
| Prematurity | 4 (0.7) | 3 (1.1) | 1 (0.3) | 0.19 |
| **SARS-CoV-2 antibodies** |  |  |  |  |
| IgG | 11 (1.9) | 6 (2.4) | 5 (1.5) | 0.43 |
| IgM | 2 (0.3) | 2 (0.8) | 0 (0.0) | 0.10 |

Fourteen children were born from women testing positive to RT-PCR COVID-19 infection the day of delivery. Their main characteristics are shown in Supplementary Table 2. Their gestations lasted between 37 weeks + 4 days and 40 weeks + 1 day. They weighted between 2755 and 3500 g at delivery. Three children were admitted in the ICU: one due to respiratory distress; he had Apgar 1’ = 4 and Apgar 5’ = 8; one child because of hypoglycaemia (Apgar 1’ = 9, Apgar 5’ = 10) and the third child because of social indication (Apgar 1’ = 9, Apgar 5’ = 10). The other 11 children were healthy, their Apgar 1’ was 9 and their Apgar 5’ ranged 9 – 10; they did not required admission in the ICU. Naso-pharyngeal swabs were obtained for RT-PCR analysis at least twice for each child: one the day of delivery and another the day after; they were all negative. A test for antibodies anti-SARS-CoV-2 was carried out; only a child tested positive for IgG and negative for IgM. The remaining 13 children were negative for both IgG and IgM.

### Strengths and limitations

In this article we are reporting the inception and first results of a cohort of women who have been pregnant in the COVID-19 pandemic in Spain and their children. The study has some limitations. Firstly, it has been designed for recruiting 1000 women and children under the assumption that prevalence of COVID-19 infection in Spain by 31^st^ March would be about 15%, as suggested by the first version of a report from the Imperial College (36). The fact that a Spanish national study later reported the prevalence to be 5% (37) could make our study underpowered. If it happens, we would deal it by enlarging our cohort. Secondly, information regarding exposure to people infected by SARS-CoV-2 or risk activities is self-reported, which makes it less reliable than recorded variables such as those regarding pregnancy control or delivery results; therefore, some information bias could be expected, although it could possibly be a non-differential one. Thirdly, women participating in the study could be more motivated than women rejecting it. In this regard, recruitment of women in sub-cohort 1 was delayed for some weeks, as many women were reluctant to come back to the hospital in the middle of the pandemic, where news about hospital activity and number of admitted or dead patients by COVID-19 were alarming. As the first wave went on, they became much more cooperative and widely agreed in participating in the study. This did not happen in sub-cohorts 2 and 3 as they were actually recruited in a routine visit to hospital. Therefore, we could not rule out the possibility of participants in sub-cohort 1 being different from participants in sub-cohorts 2 and 3. Nevertheless, participants in all three cohorts shared most characteristics as shown in Table 2, which makes such a differential participation less likely. Fifthly, antibodies anti-SARS-CoV-2 in neonates are being determined via immunochemoluminiscency, which seems to be less sensitive than ELISA. In spite of the potential losing of accuracy, we do prefer it for neonates as we would consider unethical to obtain a sample of blood by venopuncture from a neonate unless clinical reasons justify it.

The study has some strengths too. Firstly, we are recruiting women in one of the developed countries earlier and more affected by COVID-19 (38,39). Secondly, we are identifying three sub-cohorts whose higher exposure to COVID-19 would be in different pregnancy trimester, so that we could study the effect of early and late infection on both mother and child health. Thirdly, this study takes place in a country with public health system of universal coverage; therefore, differences that could be find among woman and child health are not expected to be due to differences in health care accessibility. Fourthly, we could be able to compare this cohort with a previous one recruited in the same hospital in 2018, so that we expect to measure differences in health care due to COVID-19 pandemic affecting women and children irrespective they were or not infected by SARS-CoV-2. Fifthly, the cohort being recruited in only a hospital somehow guaranties homogeneity in health care and collecting information. Further on, women in this cohort could join the International Registry of Coronavirus Exposure in Pregnancy (IRCEP, https://corona.pregistry.com) so that results from this cohort could be compared with those of women elsewhere.

## CONCLUSION

MOACC study has recruited more than 1000 pregnant women and is recruiting their neonates during the COVID-19 pandemic evolution in Spain in order to ascertain the impact COVID-19 would have on both mother and child health. Characteristics of three different sub-cohorts would allow us to study such an effect on each pregnancy trimester and to compare this cohort with a previous one recruited in the same hospital before the beginning of the pandemic, which could also allow to understand what changes have occurred in pregnancy health care during the pandemic and what effects those changes could have.

## Data Availability

Data are available upon request

## Author contributions

Conceptualization, JL, MJC; methodology, JL, CL, PG, MJC; formal analysis, JL, MFO, MJC; investigation, CL, PG, YJ, MJC; data curation, JL, MFO; writing—original draft preparation, JL; writing—review and editing, JL, CLM, MJC; funding acquisition, JL, MJC. All authors have read and agreed to the published version of the manuscript.

## Acknowledgments

The authors acknowledge the cooperation of Dr Jorge Calvo-Montes, Service of Microbiology at University Hospital Marqués de Valdecilla and Dr Marcos López-Hoyos, Service of Immunology at University Hospital Marqués de Valdecilla, for the performance of RT-PCR analyses and anti-SARS-CoV-2 antibodies determinations. The Service of Gynaecology and Obstetrics and the Service of Paediatrics at University Hospital Marqués de Valdecilla collaborated in recruiting patients.

## Data sharing

Data will be available at the repository ISCIII-COVID19 (www.isciii.es). Before that availability, data will be shared upon reasonable request to the authors.

