## Supplemental Tables 1 and 2 for "Cohort profile: COVID-19 in a cohort of pregnant women and their descendants, the MOACC-19 study"

Correspondence:

Supplementary Table 1. Characteristics of women tested positive for SARS-CoV-2 infection via RT-PCR, subcohorts 1 and 2

| Variable | Category | N (%) |
|---|---|---|
| **Study cohort** | Sub-cohort 1 (Women delivering before 26/May/2020) | 5 |
| | Sub-cohort 2 (Women delivering from 26/May/2020 on) | 9 |
| **Partner RT-PCR** | Negative | 7 |
| | Positive | 7 |
| **Age** | <25 years | 1 (7.1) |
| | 25-29 years | 3 (21.4) |
| | 30-34 years | 4 (28.6) |
| | 35-39 years | 4 (28.6) |
| | ≥40 years | 2 (14.3) |
| **Education level** | Primary studies | 3 (21.4) |
| | Secondary studies | 2 (14.3) |
| | Vocational training | 2 (14.3) |
| | University studies | 7 (50.0) |
| **Work status** | Unemployed / no active worker | 5 (35.7) |
| | Employed | 9 (64.3) |
| | Student | 0 |
| **Occupation** | Health care worker | 4 (28.6) |
| | Working in restaurant or commerce | 2 (14.3) |
| | Others | 3 (21.4) |
| **Fertilization type** | Natural | 12 (92.3) |
| | Artificial insemination | 1 (7.7) |
| | In vitro fertilization (own ovule) | 0 |
| | In vitro fertilization (ovodonation) | 0 |
| | Missing | 1 (7.7) |
| **Smoker in pregnancy** | No | 14 (100.0) |
| | Yes | 0 |
| **Alcohol consumption in pregnancy** | No | 14 (100.0) |
| | Yes | 0 |
| **Parity** | 1 | 9 (64.3) |
| | 2 | 1 /7.1) |
| | ≥3 | 4 (28.6) |
| **Type of delivery** | Eutocic | 8 (57.1) |
| | Caesarean rate | 4 (28.6) |
| | Instrumental | 2 (14.3) |
| **Age (years)** | mean±sd (min, max) | 32.9±5.1 (23 – 40) |
| **Gestational age leave work (weeks)** | mean±sd (min, max) | 23.4±12.7 (0 – 36) |
| **Pregestational BMI (kg/m$^2$)** | mean±sd (min, max) | 23.3±4.0 (18.6 – 32.0) |

| | | |
|---|---|---|
| **Gestational weight gain (kg)** | mean±sd (min, max) | 11.2±4.1 (8 – 22) |
| **Symptoms associated with COVID-19** | Fever | 3 |
| | Chills | 3 |
| | Intense tiredness | 2 |
| | Sore throat | 3 |
| | Cough | 3 |
| | Short of breathing | 2 |
| | Headache | 3 |
| | Nausea | 1 |
| | Loss of sense | 2 |

Supplementary Table 2. Characteristics of children born from women tested positive to SARS-CoV-2 infection using RT-PCR

| Variable | Category | N = 7 (%) |
|---|---|---|
| **Gestational age at birth** | **<34 weeks** | 0 |
| | **34-36 weeks** | 0 |
| | **≥37 weeks** | 14 (100.0 |
| **Birth weight** | **<2500 g** | 0 |
| | **2500-4000 g** | 14 (100.0) |
| | **>4000 g** | 0 |
| **Gender** | **Women** | 4 (28.6) |
| | **Men** | 10 (71.4) |
| **Twin or singleton** | **Singleton** | 14 (100.0) |
| | **Twin** | 0 |
| **Breastfeeding at hospital discharge** | **Exclusive breastfeeding** | 4 (28.6) |
| | **Mixed** | 6 (42.9) |
| | **Infant formula** | 4 (28.6) |
| **Neonatology admission** | **No** | 11 (78.6) |
| | **Yes** | 3 (21.4) |
| **Birth size** | **mean±sd (min, max)** | 50.0±2.4 (47 – 53) |
| **Birth cephalic perimeter** | **mean±sd (min, max)** | 34.7±1.5 (33 – 39) |
| **Apgar 1’** | **mean±sd (min, max)** | 8.4±1.4 (4 – 9) |
| **Apgar 5’** | **mean±sd (min, max)** | 9.2±0.9 (7 – 10) |
| **pH at birth** | **mean±sd (min, max)** | 7.27±0.05 (7.20 – 7.36) |
| **SARS-CoV-2 via RT-PCR** | | All negative |
